# Smartphone Camera Based Assessment of Adiposity: A Multi-Site Validation Study

**DOI:** 10.1101/2021.06.10.21258595

**Authors:** Maulik D. Majmudar, Siddhartha Chandra, Samantha Kennedy, Amit Agrawal, Mark Sippel, Prakash Ramu, Apoorv Chaudhri, Antonio Criminisi, Brooke Smith, Steven B. Heymsfield, Fatima Cody Stanford

## Abstract

**Background:** Body composition is a key component of health in both individuals and populations, and excess adiposity is associated with an increased risk of developing chronic diseases. Body mass index (BMI) and other clinical or consumer-facing tools for quantifying body fat (BF) are often inaccurate, cost-prohibitive, or cumbersome to use. The aim of the current study was to evaluate the performance of a novel automated computer vision method, visual body composition (VBC), that uses two-dimensional photographs captured via a conventional smartphone camera to estimate percentage total body fat (%BF).

**Methods:** 134 healthy adults ranging in age (21-76 years), sex (61.2% women), race (60.4% Caucasian; 23.9% Black), and body mass index (BMI, 18.5-51.6 kg/m^2^) were evaluated at two clinical sites. Each participant had %BF measured with VBC, three consumer and two professional bioimpedance analysis (BIA) systems, as well as air displacement plethysmography (ADP). %BF measured by dual-energy X-ray absorptiometry (DXA) was set as the reference against which all other estimates were compared.

**Results:** Relative to DXA, VBC had the lowest mean absolute error and standard deviation (2.34%±1.83%) compared to all other evaluated methods (p<0.05 for all comparisons). %BF measured by VBC also had very good concordance with DXA (Lin’s concordance correlation coefficient, CCC: overall 0.94; women 0.92; men 0.90); whereas BMI had very poor concordance (CCC: overall 0.45; women 0.40; men 0.74). Bland-Altman analysis of VBC revealed the tightest limits of agreement (LOA) and absence of significant bias relative to DXA (bias 0.85%, R^2^=0.01; p=0.41; LOA −4.7% to +6.4%), whereas all other evaluated methods had significant (p<0.01) bias and wider limits of agreement.

**Conclusion:** In this first validation study of a novel, accessible, and easy-to-use system, VBC body fat estimates were accurate and without significant bias compared to DXA as the reference; VBC performance exceeded those of all other BIA and ADP methods evaluated. The wide availability of smartphones suggests that the VBC method for evaluating %BF can play a major role in quantifying adiposity levels in a wide range of settings.

**TRIAL REGISTRATION:** Funded by Amazon, Inc., Seattle, WA.

## INTRODUCTION

Body composition is associated with cardiorespiratory fitness and longitudinal health outcomes^1^. In clinical practice, body composition assessment is often used to evaluate dietary habits, excess adiposity and malnutrition, weight loss following bariatric surgery, and the sarcopenia that often evolves with aging^2^. Excess adiposity impairs functional performance, is a major risk factor for developing chronic diseases, and is often accompanied by poor self-esteem^3,4,5^.The increased risk of chronic diseases that accompany excessive fat accumulation are the leading cause of death globally and contribute to an estimated $210 billion in medical costs in the US annually^6,7^.

In clinical practice, thresholds for body weight classifications are distinguished using BMI where adults with BMI ≥25 and ≥30 kg/m^2^ are characterized with overweight and obesity, respectively^8,9,10^. A limitation of BMI, however, is that it cannot discern the fat component of body mass from lean tissues. As such, adiposity levels are often misclassified in those who deviate from normalized lean mass percentages including older adults who have lost muscle with age and athletic individuals with more muscular builds^11,12^. As studies have become more inclusive^13^, it has also become apparent that body composition, specifically percent body fat (%BF), varies across race and ethnic groups even after controlling for age and BMI, which leaves placement of weight category thresholds questionable when applied to the general public^14,15,16,17^. Due to these limitations, BMI is an imperfect obesity screening tool despite its widespread clinical application^18,19,20,21^. Alternative body composition technologies, such as bioelectrical impedance analysis (BIA), calipers and anthropometric measurements, are commonly used due to time and ease of measurement at the expense of accuracy^22,23,24,25^. Imaging techniques such as dual-energy X-ray absorptiometry (DXA), computed tomography (CT), or magnetic resonance imaging (MRI) are currently considered the reference standards in body composition analysis due to their ability to discriminate and localize soft tissues^26,27^. Nevertheless, these methods are rarely applied in routine care due to concerns with cost, convenience, accessibility, radiation dose, and equipment size.

Recently, advancements in optical imaging technology have offered innovative solutions for creating accurate, precise, and relatively inexpensive methods of assessing body size, shape, and composition^28,29^. Three-dimensional imaging devices have made it possible to easily obtain thorough body measurements and estimate composition without requiring considerable skill or additional instruction^30^. However, due to their size and cost, ranging anywhere between $10,000 to $20,000 USD, current 3D optical systems remain largely unavailable to most consumers.

The gap in available accurate and inexpensive tools for consumers to estimate and track their adiposity level led us to develop a novel imaging approach for quantifying %BF. The application of machine learning, specifically deep learning, to the task of body fat estimation from 2D optical images has not previously been explored sufficiently despite widespread potential because of the inherent complexities in acquiring reliable ground truth measurements and a lack of large datasets in this domain. The aim of this study was to evaluate the performance of VBC, a novel body composition analysis system, in estimating %BF directly from 2D images captured by a conventional smartphone as compared to multiple other commercial body composition analysis methods with DXA as the reference measurement.

## METHODS

### Trial Design and Oversight

The VBC analysis system was examined in a prospective, clinical validation study conducted at two clinical trial sites: Massachusetts General Hospital (MGH), and Pennington Biomedical Research Center (PBRC), Louisiana State University. The study protocol was approved by the Advarra Institutional Review Board (Columbia, MD) as well as the MGH and PBRC Institutional Review Boards.

Participants were contacted by a recruiter who performed pre-screening based on demographic information as well as inclusion and exclusion criteria. Eligible participants were asked to arrive at their respective facility for a single 2–3-hour visit following a 4-hour fast. Upon arrival, they were provided a copy of the consent form and a private room for the consenting process. Those who agreed to participate completed the following assessments for %BF: DXA and VBC scans, three consumer-grade bioimpedance analysis (cBIA) smart scale evaluations, two professional BIA (pBIA) system evaluations, and air displacement plethysmography (ADP). Women with reproductive potential also completed a urine pregnancy test prior to undergoing these assessments.

### Trial Participants

Participants were healthy adults recruited using web-based questionnaires, direct phone calls, media, and community outreach. Included men and women were generally in good health, between the ages of 21 and 80 years, weighed less than 400 lbs (181 kg), and willing to comply with study procedures. Potential participants were excluded if they had medical implants such as a pacemaker or a total knee replacement or had previously undergone body altering procedures such as arm or leg prosthesis, amputation, or breast augmentation. Participants were also excluded if they took loop diuretics within 6 hours of their scheduled visit, had a diagnosis of heart failure, or were undergoing active cancer treatment.

### Trial Procedures

For each of the participants, trained facility staff acquired the following data: demographic information such as age, sex, ethnicity, height, and weight; anthropometric measurements taken at the waist, hip, arm, and thigh; 2D photographs captured by a smartphone camera; %BF estimates from consumer and professional BIA scales, ADP, and DXA; only participants at PBRC underwent ADP.

### Anthropometry

Circumference measurements were taken at the waist, hip, arm, and thigh by trained staff at conventional anatomic locations. Measurements were recorded in centimeters.

### VBC scan

Participants were dressed in minimal, form-fitting clothing (**Figure 1**) without socks, shoes, or any protruding wearables (watches, jewelry, etc.), such that the mid-thigh and belly button areas were visible to the smartphone camera. Each participant was asked to stand in an “A” pose and then had four photographs (front, back, left-side, and right-side profiles) taken with an iPhone-10 (Apple, Inc.) front-facing camera with their faces out of frame.

**Figure 1.**
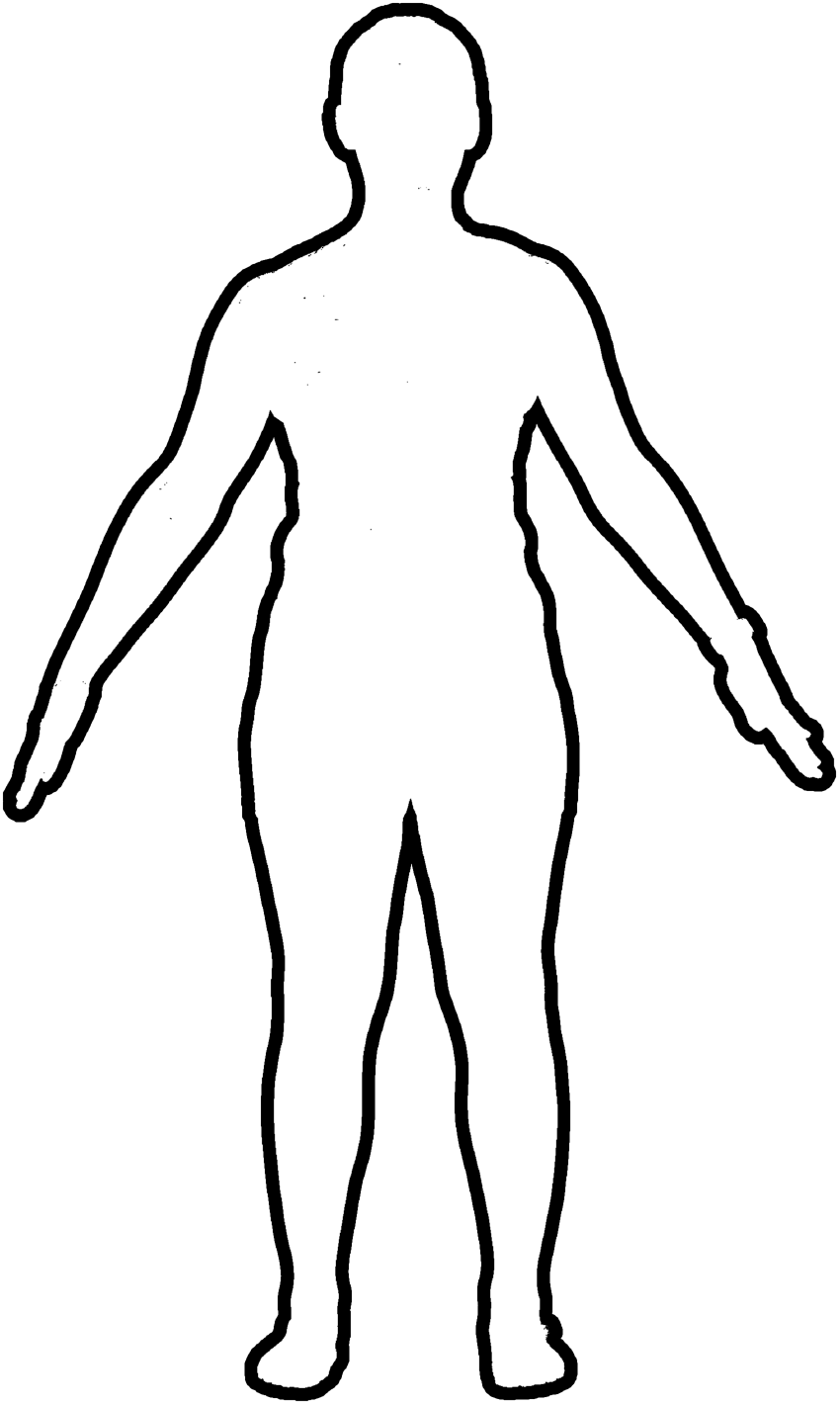
VBC’s computer vision-based algorithm requires images of the user’s front and back while holding an “A” pose as inputs to the model.

#### Computer Vision Model

The body composition estimation algorithm consists of a bespoke convolutional neural network (CNN)^31,32^ that was trained to estimate %BF directly from two input photographs (front and back) of the user standing in an A pose as shown in **Figure 1**. The algorithm does not need accurate 3D scans nor high quality images; the photos were acquired through a smartphone positioned 4-6 feet away from the participant above their knee height. Of note, the side images are used to generate a three-dimensional body model (using a different computer vision algorithm), which is a feature of the commercial product (Amazon Halo); but those images are not used for estimation of %BF. The VBC %BF algorithm was developed using the Python programming language (Python Software Foundation; available at www.python.org) and uses the PyTorch machine learning framework (available at www.pytorch.org and maintained by Facebook) for training and evaluating the CNN. The developed model was trained on GPU-enabled machines for speed.

#### Training

The training dataset is a separate repository consisting of front and back photos of participants taken from a smartphone, associated with %BF ground truth. A CNN model was first trained to delineate the body in the image and remove the background pixels with a high degree of precision. After background removal, the front and back photos are normalized to a canonical size to account for variations in camera distance. Using the normalized front and back photos as inputs for training, a second CNN model is pre-trained to analyze overall shape and details of the body from 2D images and automatically extract discriminative visual features relevant to body composition. The architecture of this model uses multiple convolutional blocks with additional branches for multi-scale feature extraction. The multi-scale extension allows the network to utilize finer/higher resolution image features and helps in capturing details across the body fat spectrum. This model is trained to be resilient to noise in the input images, robust to normal variations in illumination and camera orientation, and to be able to work across different camera devices and color-spaces (grayscale/RGB). Next, transfer learning is applied to fine-tune this model using DXA %BF data. Note that the data from the current study was not used to train, fine-tune or internally validate these models.

#### Runtime

At runtime, a pair of front and back images are sent into the CNN and the output is a single %BF measurement.

### Dual-Energy X-ray absorptiometry

Total body fat was measured on each participant with a Hologic Discovery A or Hologic Horizon A DXA system (Hologic, Inc., Marlborough, MA, USA). Both DXA systems were calibrated and operated according to manufacturer guidelines. Attired in minimal clothing, participants were asked to lay flat on the DXA table for about 10 minutes while the device performed the scan. All scans were evaluated with Hologic Apex software version 5.6 and the National Health and Nutrition Examination Survey (NHANES) Body Composition Analysis calibration feature was disabled.

### Bioimpedance Analysis

Three consumer weight scales capable of BIA-based body composition analysis were included in the protocol: FitBit Aria 2 (Fitbit, San Francisco, CA); Tanita BF-684W (Tanita, Tokyo, Japan); and Renpho ES-24M-W/B (Joicom Corporation, Anaheim, CA). These scales are designated as cBIA 1, cBIA 2, and cBIA 3, respectively, in the sections that follow. Participants were weighed in duplicate on the consumer scales, and the results were averaged for analyses. All participants also underwent professional BIA (pBIA) at PBRC with an InBody S10 (InBody Co., Seoul, Korea) and at MGH with a RJL system (Quantum IV, RJL Systems, Clinton Township, MI, USA). These are designated as pBIA1 and pBIA 2, respectively, in the sections that follow, and were analyzed separately. Both InBody S10 and RJL Quantum IV use a tetrapolar 8-point tactile electrode system. The device measures impedance, resistance, and reactance in body segments at multiple frequencies. Each participant was measured once following cleaning of the electrodes with alcohol.

### Air Displacement Plethysmography

Participants who were evaluated at PBRC also had %BF assessed with the BOD POD ADP device (BodPod Gold Standard Body Composition Tracking System, COSMED, Rome, Italy). In addition to the specific form fitted clothing for this study, participants put on a swim cap before entering the device. The BOD POD body composition test was performed once with each evaluation including two measurements of body volume that were averaged and then corrected for thoracic gas volume using the system software (v4.5.0). Fat mass and %BF were calculated from body density by BOD POD software using Siri’s equation^33^.

### Statistical Methods

Descriptive statistics were computed for the participant characteristics stratified by sex, where appropriate. Fixed bias (or mean error) was calculated as the difference between %BFDXA and %BF estimates from all other methods evaluated: VBC, cBIA1-3, pBIA1-2, and ADP. Mean absolute error (MAE), standard deviation (SD) of absolute error, and mean absolute percent error (MAPE) were calculated for all %BF estimates and stratified by sex, BMI, and race. Wilcoxon signed rank test was used to compare matched samples to assess whether their population mean ranks differ (i.e., paired difference test) for the overall study population and stratified by sex. Pearson correlation and Lin’s concordance correlation coefficient (CCC) between DXA and all other methods were also calculated, and stratified by sex. The method of Meng et al^34^ was used to determine whether VBC was significantly better correlated to the criterion method of DXA compared to the cBIA 1-3, pBIA 1-2, and ADP measurements. Bland-Altman plots were created to determine the mean difference and 95% limits of agreement (LOA) between DXA reference standard and VBC as well as all other methods. All analyses were conducted using Microsoft Excel (Microsoft, Inc., Redmond, WA) and Python. Significance was set at an alpha level of 0.05, 2-tailed.

## RESULTS

### Participants

A total of 406 adults were initially screened for this study. Of those, 199 met all inclusion and exclusion criteria and were considered eligible. 138 participants were enrolled into the clinical study and 134 participants were included in the final analysis; four participants (2.9%) were removed from the final analysis due to poor image quality (**Figure 2**). The demographic and anthropometric characteristics of the final study sample are shown in **Table 1**. The ethnic and racially diverse sample was 60.4% Caucasian, 23.9% Black, 6.7% Asian, 3.0% Hispanic, 0.7% American Indian and the remaining 5.2% Multiracial, across the two study sites. Participant’s mean age was 43±14.7 years (range, 21-76 years) and BMI 29.7±6.5 kg/m^2^ (range, 18.5-51.6 kg/m^2^). DXA-measured %BF was 39.4±7.2% in women and 28.6±6.4% in men.

**Table 1.** Subject characteristics

|  | *All* | *Males* | *Females* |
| --- | --- | --- | --- |
| **Total number** | 134 | 52 | 82 |
| Caucasian (%) | 81 (60.4) |  |  |
| Black (%) | 32 (23.9) |  |  |
| Asian (%) | 9 (6.7) |  |  |
| Hispanic (%) | 4 (3.0) |  |  |
| American Indian (%) | 1 (0.7) |  |  |
| Multiracial (%) | 7 (5.2) |  |  |
| **Age (years)** | 43 ± 14.7 |  |  |
| **Height (cm)** | 167.8 ± 10.2 |  |  |
| **Weight (kg)** | 84.0 ± 20.8 |  |  |
| **BMI (kg/m<sup>2</sup>)** | 29.7 ± 6.5 |  |  |
| **Waist circumference (cm)** |  | 102.2 ± 14.3 | 96.2 ± 16.3 |
| **Hip circumference (cm)** |  | 108.2 ± 11.5 | 110.1 ± 14.3 |
| **Weight-to-hip ratio** |  | 0.94 ± 0.08 | 0.87 ± 0.07 |
| **DXA %BF** |  | 28.6 ± 6.4 | 39.4 ± 7.2 |
| Abbreviations: BMI, body mass index; DXA, dual-energy X-ray absorptiometry; %BF, percent body fat. Results are X ± s.d. |  |  |  |

**Figure 2.**
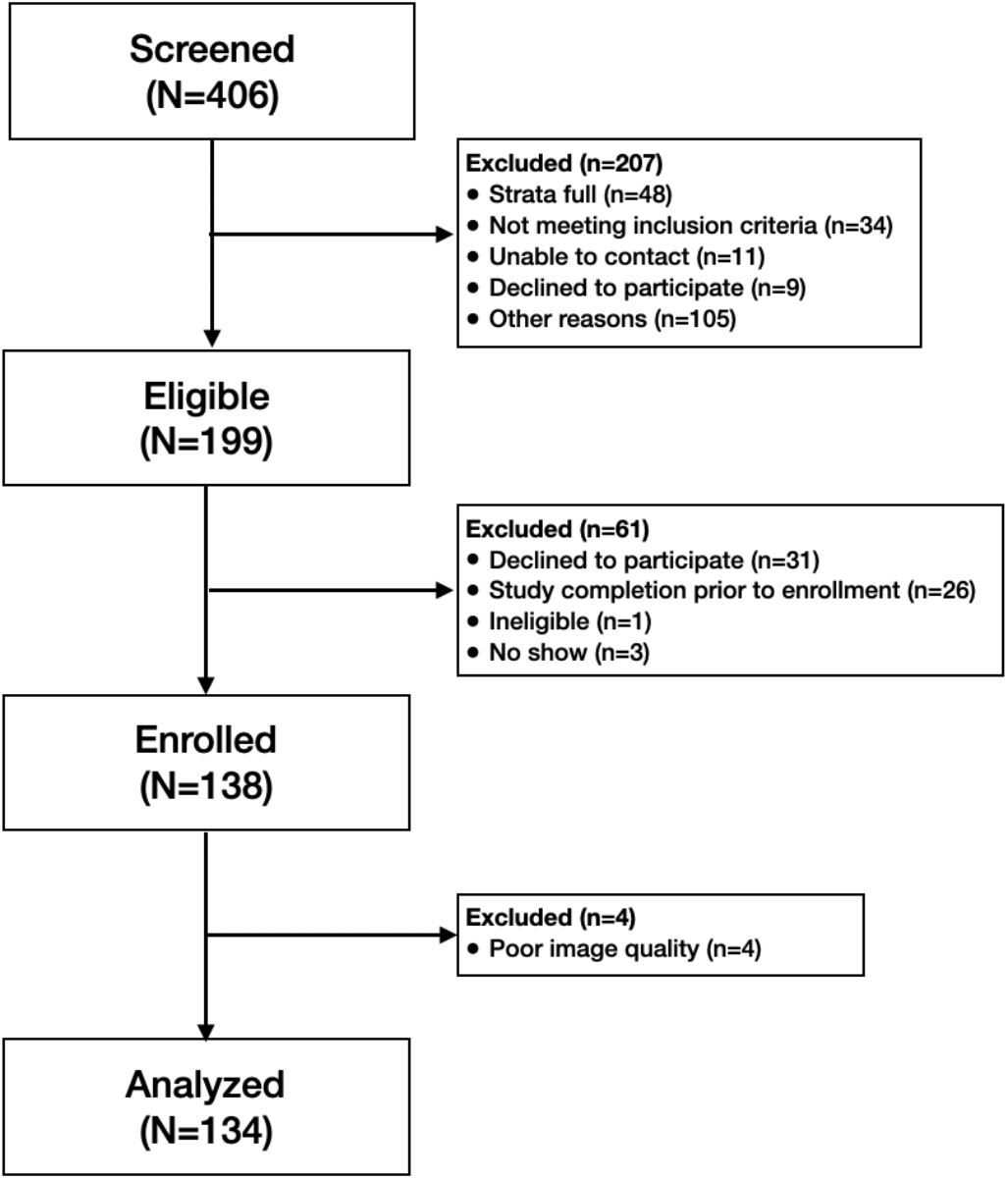
Consort Diagram.

### Body Composition

VBC achieved the lowest error in estimating %BF with MAE and SD of 2.34%±1.83% and MAPE of 7.2% compared to DXA, with an overall bias of 0.85%. cBIA 1, 2, and 3 had bias of −0.67%, −0.12%, and −2.93%, respectively. MAE and SD for these three devices were 4.48%±4.01%, 4.91%±8.7%, and 5.85%±4.86%, respectively. The bias, MAE, and SD of the pBIA 1, pBIA 2, and ADP systems were −1.07%, 3.13%±2.10%; 0.64%, 4.72%±3.0%; and 0.55%, 3.14%±2.24, respectively. The key performance measures, including overall bias, MAE, SD, and concordance correlation coefficient (CCC) of DXA as compared to the seven devices evaluated are presented in **Table 2**. Compared to DXA, VBC demonstrated very high concordance (CCC=0.94) in the overall sample, which was higher than all other methods evaluated, including ADP (**Table 2**).

**Table 2.** Comparison of %BF estimates to DXA references stratified by sex.

| **Table 2.** Comparison of %BF estimates to DXA references stratified by sex. |  |  |  |  |  |  |  |
| --- | --- | --- | --- | --- | --- | --- | --- |
|  | **VBC** | **cBIA 1** | **cBIA 2** | **cBIA 3** | **pBIA 1** | **pBIA 2** | **ADP** |
| **All** |  |  |  |  |  |  |  |
| Bias (%) | 0.85 | -0.67 | -0.12 | -2.93 | -1.07 | 0.64 | 0.55 |
| MAE (%) | 2.34 ± 1.8 | 4.48 ± 4.0* | 4.91 ± 8.7* | 5.85 ± 4.9* | 3.13 ± 2.1* | 4.72 ± 3.0* | 3.14 ± 2.2* |
| MAPE (%) | 7.2 | 14.2 | 15.4 | 16.7 | 10.3 | 14.0 | 9.7 |
| CCC | 0.94 | 0.80* | 0.58* | 0.72* | 0.92 | 0.87* | 0.92 |
| **Men** |  |  |  |  |  |  |  |
| Bias (%) | 1.21 | 0.22 | 1.55 | -1.45 | -1.41 | -4.58 | 2.24 |
| MAE (%) | 2.23 ± 1.8 | 4.53 ± 5.0* | 6.23 ± 13.4* | 4.11 ± 3.3* | 3.37 ± 2.4 | 5.01 ± 3.0* | 3.50 ± 2.4 |
| MAPE (%) | 8.4 | 18.2 | 22.6 | 15.5 | 13.8 | 17.4 | 12.5 |
| CCC | 0.90 | 0.57* | 0.29* | 0.74* | 0.87 | 0.54* | 0.88 |
| **Women** |  |  |  |  |  |  |  |
| Bias (%) | 0.62 | -1.20 | -1.15 | -3.82 | -0.85 | 3.99 | -0.51 |
| MAE (%) | 2.41 ± 1.8 | 4.45 ± 3.3* | 4.10 ± 3.1* | 6.89 ± 5.3* | 2.98 ± 1.8* | 4.54 ± 3.0* | 2.91 ± 2.1 |
| MAPE (%) | 6.4 | 11.8 | 10.9 | 17.4 | 8.2 | 11.9 | 8.0 |
| CCC | 0.92 | 0.78* | 0.79* | 0.62* | 0.90 | 0.79* | 0.91 |
| Abbreviations: ADP, air displacement plethysmography; cBIA, consumer bio-impedance analysis; CCC, concordance correlation coefficient; MAE, mean absolute error; MAPE, mean absolute percent error; pBIA, professional bio-impedance analysis; VBC, visual body composition. *$P < 0.05$ in comparison to VBC-DXA. Results are $X \pm s.d.$ | | | | | | | |

Further sub-cohort analyses of the performance of all devices evaluated for estimating %BF classified by sex, BMI, and ethnicity are summarized in **Table 2 and 3**. When stratified by sex, VBC continues to show the lowest MAE and MAPE values. VBC has MAE±SD 2.23%±1.84%, MAPE 8.4% in men and MAE±SD 2.41%±1.81%, MAPE 6.4% in women. VBC also had very good concordance for both women (CCC=0.92) and men (CCC=0.90), as shown in **Table 2**.

**Table 3.** Comparison of %BF estimates to DXA references stratified by BMI and ethnicity.

|  |  |  |  |  |  |  |  |
| --- | --- | --- | --- | --- | --- | --- | --- |
|  | **VBC** | **cBIA 1** | **cBIA 2** | **cBIA 3** | **pBIA 1** | **pBIA 2** | **ADP** |
| **BMI<25 kg/m<sup>2</sup>** |  |  |  |  |  |  |  |
| Bias (%) | 0.94 | -1.03 | 0.76 | -5.05 | -3.34 | 0.83 | -3.00 |
| MAE (%) | 2.04 ± 1.3 | 6.36 ± 3.5 | 5.75 ± 3.3 | 7.66 ± 4.6 | 4.29 ± 2.2 | 4.41 ± 2.8 | 3.31 ± 2.2 |
| MAPE (%) | 6.9 | 23.0 | 19.6 | 24.5 | 15.7 | 16.8 | 11.9 |
| **BMI 25-29.9 kg/m<sup>2</sup>** |  |  |  |  |  |  |  |
| Bias (%) | 0.94 | -1.03 | 0.76 | -5.05 | -1.90 | -0.70 | 0.69 |
| MAE (%) | 2.24 ± 1.9 | 3.02 ± 2.3 | 5.04 ± 14.3 | 6.00 ± 4.9 | 2.49 ± 1.4 | 3.82 ± 3.1 | 2.3 ± 1.9 |
| MAPE (%) | 8.1 | 10.1 | 17.0 | 17.3 | 8.3 | 12.9 | 7.5 |
| **BMI≥30 kg/m<sup>2</sup>** |  |  |  |  |  |  |  |
| Bias (%) | 1.75 | 1.48 | 1.40 | 1.31 | 1.62 | 1.68 | 3.58 |
| MAE (%) | 2.45 ± 2.0 | 4.44 ± 5.0 | 4.31 ± 3.6 | 4.60 ± 4.6 | 2.63 ± 2.0 | 5.58 ± 2.6 | 3.67 ± 2.3 |
| MAPE (%) | 6.6 | 11.9 | 11.6 | 11.2 | 7.3 | 14.0 | 9.6 |
| **Caucasian** |  |  |  |  |  |  |  |
| Bias (%) | 0.64 | -2.07 | -0.90 | -4.02 | -1.39 | -0.3 | 0.18 |
| MAE (%) | 2.21 ± 1.8 | 4.23 ± 3.3 | 4.99 ± 10.8 | 5.38 ± 4.8 | 3.16 ± 2.0 | 3.86 ± 2.6 | 2.77 ± 1.9 |
| MAPE (%) | 7.0 | 13.8 | 15.8 | 16.7 | 10.9 | 11.3 | 9.0 |
| **Black** |  |  |  |  |  |  |  |
| Bias (%) | 1.26 | 3.10 | 2.27 | 0.45 | 0.77 | 2.96 | 2.4 |
| MAE (%) | 2.46 ± 1.9 | 5.01 ± 5.1 | 4.25 ± 3.1 | 5.87 ± 4.2 | 3.33 ± 2.0 | 5.36 ± 3.0 | 4.48 ± 3.0 |
| MAPE (%) | 7.6 | 15.5 | 12.9 | 16 | 9.0 | 17.0 | 12.5 |
| **All others** |  |  |  |  |  |  |  |
| Bias (%) | 1.03 | -1.43 | -0.85 | -4.05 | -2.61 | 0.07 | -0.72 |
| MAE (%) | 2.36 ± 2.2 | 4.59 ± 4.1 | 5.66 ± 4.6 | 5.90 ± 6.0 | 2.61 ± 2.4 | 6.15 ± 3.1 | 2.82 ± 1.7 |
| MAPE (%) | 7.2 | 13.7 | 17.5 | 17.8 | 9.2 | 17.2 | 8.6 |
| Abbreviations: ADP, air displacement plethysmography; cBIA, consumer bio-impedance analysis; CCC, concordance correlation coefficient; MAE, mean absolute error; MAPE, mean absolute percent error; pBIA, professional bio-impedance analysis; VBC, visual body composition. Results are X ± s.d. |  |  |  |  |  |  |  |

Table 3 illustrates results stratified by BMI. Once again, in all three BMI categories VBC achieves the lowest MAE and MAPE values. BMI<25kg/m^2^ MAE±SD 2.04%±1.33%, MAPE 6.9%. BMI 25-29.9kg/m^2^ MAE±SD 2.24%±1.92%, MAPE 8.1%. BMI≥30kg/m^2^ MAE±SD 2.45%±1.99%, MAPE 6.6%. **Table 3** also illustrates results stratified by race and ethnicity. VBC continues to show the lowest MAE and MAPE errors out of all methods compared in this study. Caucasian MAE±SD 2.21%±1.75%, MAPE 7.0%. Black MAE±SD 2.46%±1.93%, MAPE 7.6%. All others MAE±SD 2.36%±2.24%, MAPE 7.2%.

As shown in **Figure 3A** VBC achieved the lowest overall mean absolute error in estimating %BF, which was statistically significantly better than all other methods evaluated (p<0.05 for all methods), with cBIA 3 yielding the highest error. Furthermore, **Figure 3B** shows a pseudo-colored representation of the mean absolute error, both overall and stratified by sex, BMI, and ethnicity (blue indicates low error and yellow indicates high error).

**Figure 3.**
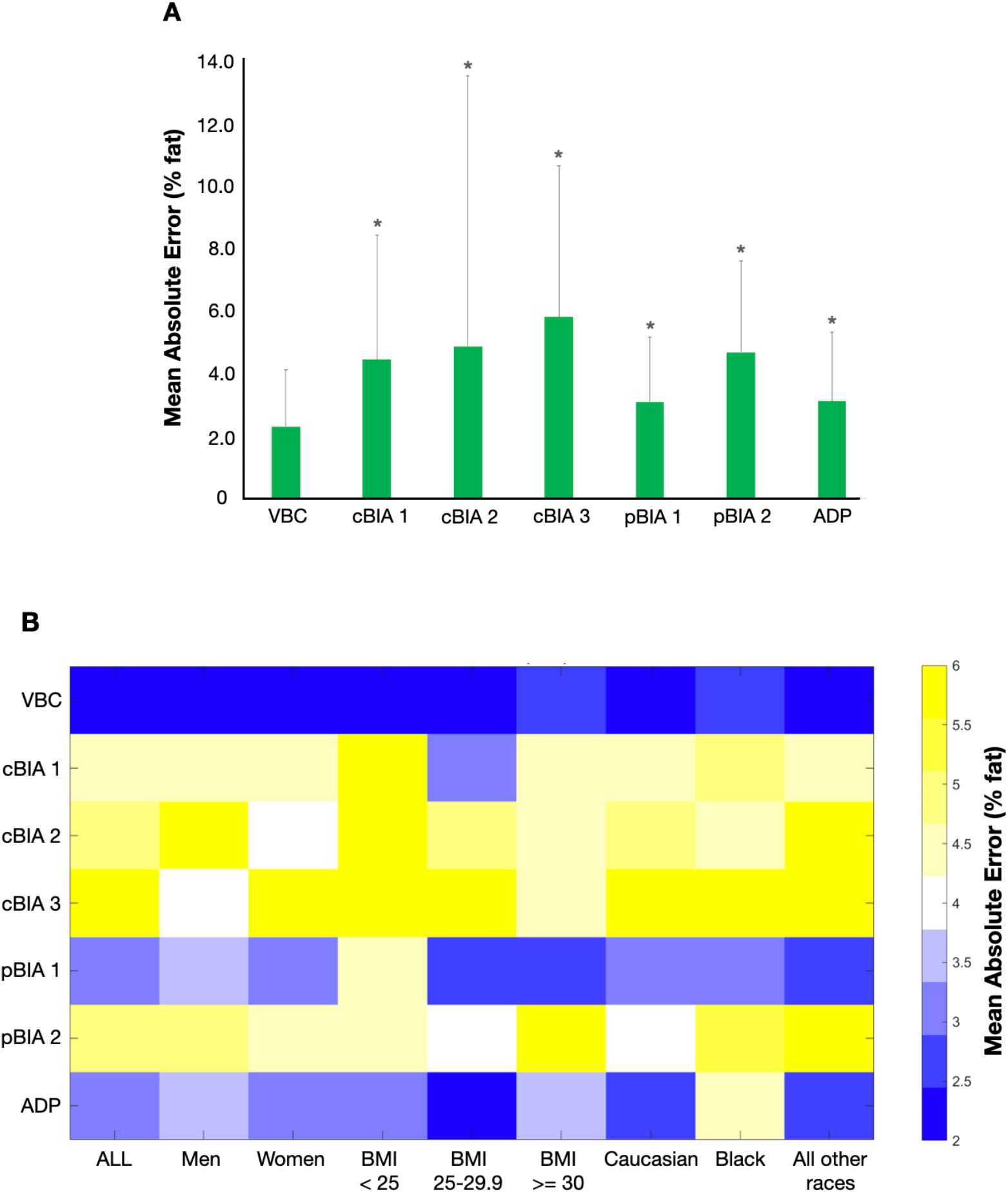
Mean absolute errors (MAE) of the various methods evaluated with DXA as the reference (3A). MAE in comparison to DXA across various methods evaluated stratified by sex, BMI, and ethnicity (3B). We defined an acceptable error range as ≤ 3% (blue). Light blue, white, and yellow shadings indicate errors outside of this range. *p<0.05 in comparison to VBC-DXA MAE.

Correlations between %BF evaluated by VBC and DXA among men and women are shown through scatter plots in **Figures 4A and 4B**, respectively. VBC achieved very good correlation for both men and women participants ((R^2^=0.85 for both sex). The corresponding Bland-Altman plot for VBC is presented in **Figure 4C** together with its limits of agreement (−4.7%, 6.4%). Individual level validity for all other methods is presented using Bland-Altman plots in **Figures 5A-5F**. VBC achieves the tightest limits of agreement without any statistically significant bias, whereas all other methods had significant bias (p<0.05) and wider limits of agreement.

**Figure 4.**
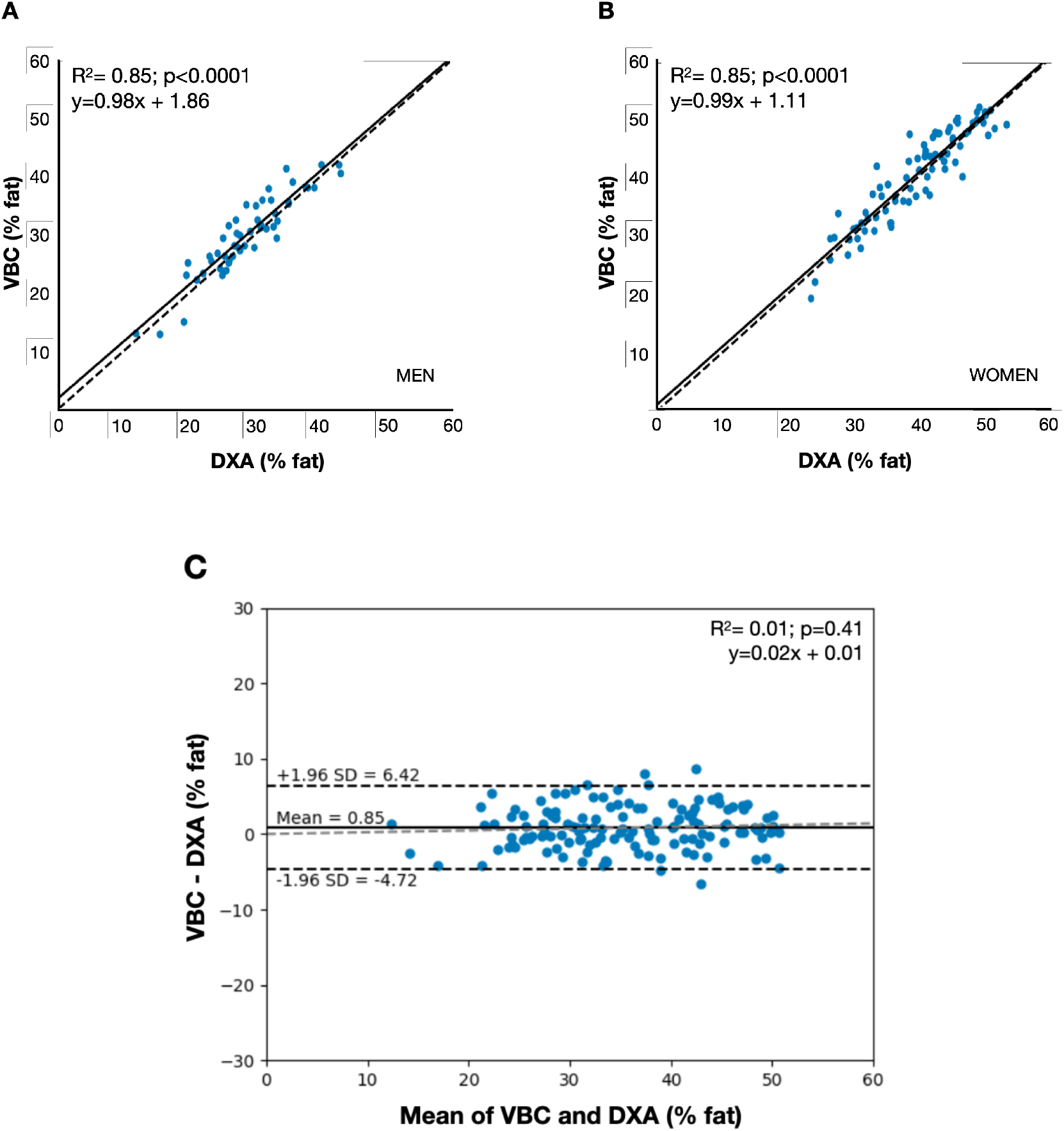
Correlation between %BF by VBC and DXA (4A men and 4B women). The dashed line is identity and the solid line is the automatically fitted regression line. The correlations in both figures are significant at p<0.0001. Bland-Altman analyses of the difference between %BF by VBC and DXA (4C). The horizontal black lines are at the mean±1.96 SD and the dashed gray lines are the fitted regression lines described by the equation in the panel.

**Figure 5.**
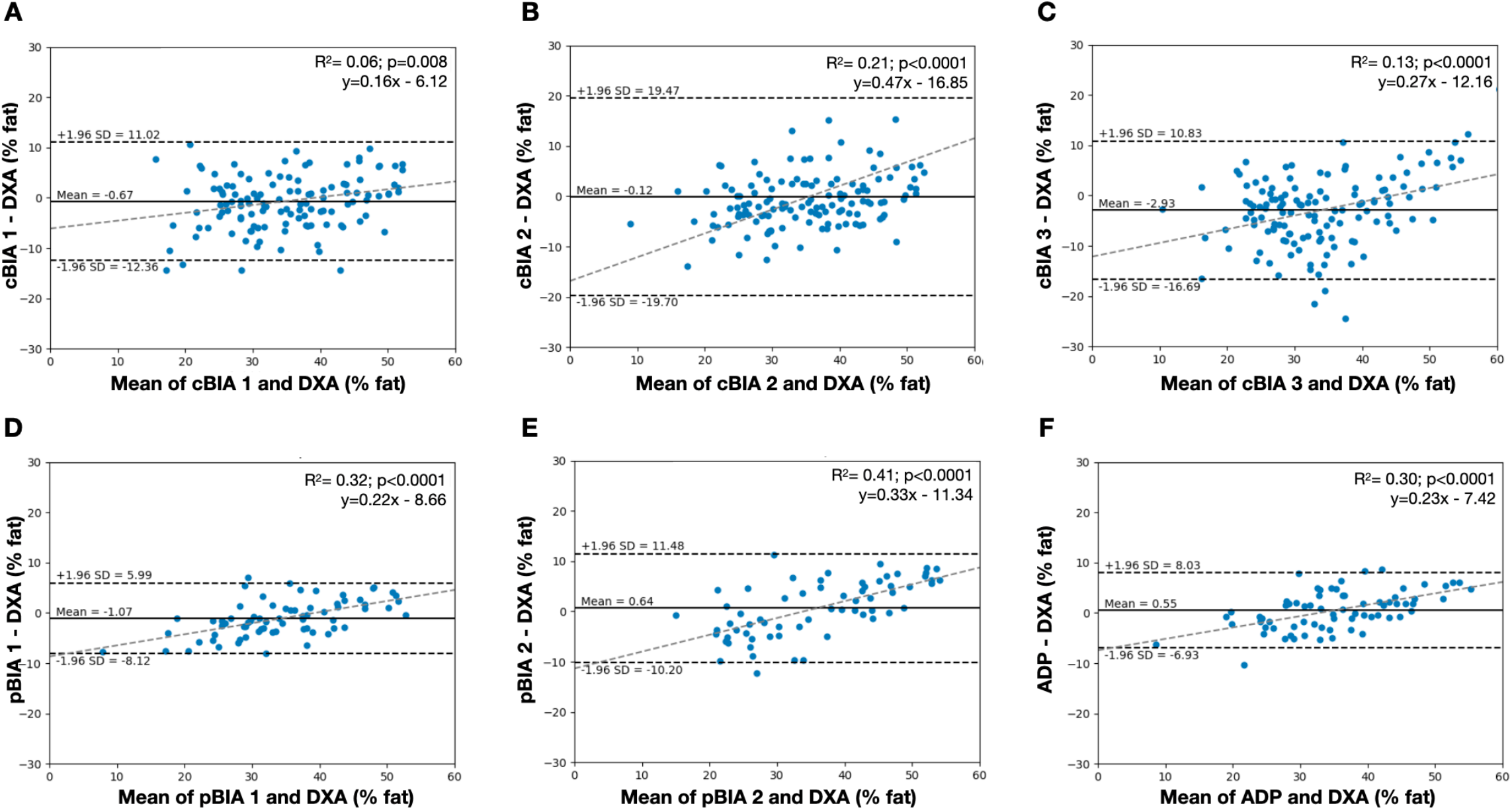
Bland-Altman analyses of the differences between %BF by DXA and the six methods evaluated for estimation of %BF. Panel A, cBIA 1; Panel B, cBIA 2; Panel C, cBIA 3; Panel D, pBIA 1; Panel E, pBIA 2; Panel F, ADP. The horizontal black lines are at the mean±1.96 SD and the dashed gray lines are the fitted regression lines described by the equations in each panel.

## DISCUSSION

There is a need for an accurate, easy-to-use, and widely accessible tool for assessment of body composition outside of specialized research facilities. The current study evaluated the performance of a novel computer-vision based model for estimating %BF from 2D photographs captured via smartphone cameras. Our findings strongly support the validity of VBC in estimating adiposity relative to DXA, the reference in this study for %BF. VBC had the lowest MAE (2.34%±1.83%), highest overall concordance with DXA (CCC, 0.95), and the tightest limits of agreement (LOA, −4.7%-6.4%) among the evaluated devices including several BIA systems and ADP.

While multiple other devices are available for capturing a person’s image and transforming the quantified information into an estimate of body composition^35,36^ VBC needs only two photographs of the participant captured via a conventional smartphone camera. These two images are securely sent to the cloud where they are processed via two different computer vision models. The first CNN model accurately delineates the person’s body and removes background pixels with a high degree of precision. The front and back photos are then size-normalized and used as input to a second CNN model tasked with analyzing the overall shape and body details. The CNN automatically extracts visual features relevant to body composition and then generates an estimate of %BF. Carletti et al described a similar framework to directly estimate %BF from depth images^37^. In contrast, VBC does not require specialized or expensive equipment like depth cameras, but instead, works with conventional smartphone cameras, making it an accessible tool at the consumer level.

Smartphones that usually include cameras are widely utilized, with over 2.5 billion users worldwide^38^. As such, there is a potential for the novel VBC tool to have wide scale use to better quantify and monitor adipose stores in persons across weight classes. Given its ease of use and low-cost, people can readily measure their body fat; for instance, biweekly or monthly, and correlate its temporal trend with their lifestyle habits, such as physical activity, dietary changes, and sleep patterns.

The VBC tool outperformed the BMI and the other evaluated %BF measuring devices examined in the current study. Body mass index has well-established limitations as a phenotypic marker of adiposity and the relatively weak associations with %BF (e.g. two people with same height and weight but different %BF would have the same BMI) were again demonstrated in the current study. Similarly, VBC outperformed commercial single frequency BIA systems for home use as they only capture the leg-leg electrical pathway and are known to have limited accuracy due to several factors that include variable participant hydration and use of population-specific %BF prediction equations^39,40^. The evaluated multi-frequency whole-body pBIA systems overcome some of the limitations present in the cBIA devices, although VBC still outperformed them both. The Bod Pod ADP device evaluated at the PBRC site is a recognized reference method for some types of studies, notably those in which radiation exposure is a concern, and at centers without available DXA systems^41^. As with the other evaluated devices, VBC similarly outperformed Bod Pod relative to DXA as the reference for %BF in the current study. Hence, given these initial findings, the VBC method appears to function at least on par, if not better, than professional systems such as pBIA and ADP.

## Limitations

While VBC performed well in the current study, several limitations of the device and our study should be noted. The CNN model was trained with photos of people wearing minimal and form-fitting clothing. Wearing full length sleeves, pants, shorts covering parts of the stomach, abdomen or thighs, or loose clothing may yield inaccurate results. Extremely dark or bright images can hide important visual information and reduce the model accuracy. Other variables that may cause inaccuracies are extreme camera tilt, camera positioned too far from the participant, holding the belly in, scanning after a large meal or an intense workout, flexing muscles or large deviations from the A pose. The model does not generate %BF estimates above 64%. The model produces a single number for %BF estimation, but currently does not provide any details on fat localization. For instance, it does not differentiate between visceral and subcutaneous adipose tissue.

The study was limited to 134 participants and a larger and more diverse sample may have further strengthened study findings. However, the study did have enough power to reach statistical significance for the primary outcome of evaluating the performance of VBC and various other methods against DXA as the reference standard.

## Conclusions

This study presents the first validation of a novel, accessible, and easy-to-use system for estimating an individual’s total body fat using only two photographs taken with a conventional smartphone. The VBC method had the lowest mean absolute error and standard deviation and the tightest limits of agreement when compared to six other state of the art methods. Percent fat estimated by VBC also had stronger concordance with those by DXA compared to the other methods and BMI. No significant bias was present for VBC relative to DXA according to a Bland-Altman analysis. These results strongly support the use and feasibility of VBC for at-home measurement and monitoring of adiposity.

## Data Availability

Data is not available for public use.

## Author Contributions

Dr. Majmudar had full access to all the data and takes responsibility for the integrity of the data and the accuracy of the data analysis. All authors comply with the International Committee of Medical Journal Editors (ICMJE) criteria for authorship of this manuscript and have approved the final version to be published.

## Data Sharing Statement

Data is not available for public use.

